## Supplemental Tables 1-7 for "Causes, characteristics, and patterns of prolonged unplanned school closures prior to the COVID-19 pandemic – United States, 2011 – 2019"

S1 Table. Cumulative incidence of prolonged unplanned school closure (PUSC) per 100 schools, United States, 2011–2019^a^.

|  | **Total Schools Closed,**  **n (cumulative incidence)** | **Unique Schools Closed,**  **n (cumulative incidence)** | **Number of all K-12 schools**^d^ |
| --- | --- | --- | --- |
| **Total** | 22,112 (18.7) | 19,582 (16.6) | 118,153 |
| **Grade level^e^** |  |  |  |
| Elementary school | 9,203 (22.9) | 7,958 (19.8) | 40,174 |
| Elementary to middle school | 4,405 (14.8) | 4,043 (13.5) | 29,853 |
| Elementary to high school | 584 (6.4) | 546 (6.0) | 9,133 |
| Middle school | 3,072 (24.2) | 2,683 (21.1) | 12,703 |
| Middle to high school | 1,021 (14.3) | 946 (13.3) | 7,129 |
| High school | 3,638 (20.5) | 3,220 (18.1) | 17,788 |
| **School Type** |  |  |  |
| Public | 21,784 (22.9) | 19,254 (20.3) | 95,045 |
| Private | 328 (1.4) | 328 (1.4) | 23,109 |
| **Urbanicity^f^** |  |  |  |
| City | 7,682 (22.7) | 7,294 (21.6) | 33,793 |
| Suburban | 7,402 (19.3) | 6,721 (17.5) | 38,354 |
| Town | 2,089 (14.3) | 1,658 (11.3) | 14,649 |
| Rural | 4,877 (15.6) | 3,847 (12.3) | 31,277 |
| **HHS region^g^** |  |  |  |
| HHS 1 | 1,021 (16.9) | 856 (14.2) | 6,029 |
| HHS 2 | 3,150 (32.7) | 3,124 (32.4) | 9,628 |
| HHS 3 | 3,109 (30.1) | 2,352 (22.8) | 10,313 |
| HHS 4 | 6,944 (35.2) | 5,852(29.6) | 19,738 |
| HHS 5 | 1,643 (7.5) | 1,638 (7.5) | 21,825 |
| HHS 6 | 3,163 (20.0) | 2,886 (18.3) | 15,796 |
| HHS 7 | 418 (6.0) | 358 (5.1) | 6,996 |
| HHS 8 | 20 (0.3) | 19 (0.3) | 5,804 |
| HHS 9 | 1,614 (9.9) | 1,561 (9.6) | 16,262 |
| HHS 10 | 1,030 (17.9) | 936 (16.2) | 5,762 |

^a^ PUSC is defined as a school closure lasting ≥5 school days, excluding any scheduled days off. Cumulative incidence of PUSC per 100 schools was computed by dividing the number of schools with PUSCs during the study period (2011-2019) by the total number of all K-12 schools and then multiplying by 100.

^b^ Schools were counted each time they experienced a PUSC across the 8-year study period.

^c^ Schools with multiple PUSCs were counted only once across the 8-year study period.

^d^ The total number of K-12 schools was obtained from the National Center for Education Statistics (NCES) by summing the number of schools reported for each academic year (2011-2012 through 2018-2019) and dividing by eight.

^e^ Grade span was not specified for 189 PUSCs, across 186 unique schools.

^f^ Urbanicity was not specified for 62 schools.

^g^ Regions of the United States Department of Health & Human Services (HHS). <https://www.hhs.gov/about/agencies/regional-offices/index.html>

S2 Table. Unique schools by the number of prolonged unplanned school closure (PUSC) events experienced, United States, 2011–2019^a, b^

|  | Unique Schools Closed | Number of Unique Schools Closed with: | | | | |
| --- | --- | --- | --- | --- | --- | --- |
|  |  | Single PUSC | Multiple PUSCs | | | |
|  |  |  | 2 PUSCs | 3 PUSCs | 4 PUSCs | Total (2-4 PUSCs) |
| Total, n (row %) | 19,582 | 17,462 (89.2) | 1,770 (9.0) | 290 (1.5) | 60 (0.0) | 2,120 (10.8) |
| HHS region^c^, n (column %) |  |  |  |  |  |  |
| HHS 1 | 856 (4.4) | 695 (4.0) | 157 (8.9) | 4 (1.4) | 0 (0.0) | 161 (7.6) |
| HHS 2 | 3,124 (16.0) | 3,098 (17.7) | 26 (1.5) | 0 (0.0) | 0 (0.0) | 26 (1.2) |
| HHS 3 | 2,352 (12.0) | 1,805 (10.3) | 355 (20.1) | 174 (60.0) | 18 (30.0) | 547 (25.8) |
| HHS 4 | 5,852 (29.9) | 4,951 (28.4) | 752 (42.5) | 107 (36.9) | 42 (70.0) | 901 (42.5) |
| HHS 5 | 1,638 (8.4) | 1,633 (9.4) | 5 (0.3) | 0 (0.0) | 0 (0.0) | 5 (0.2) |
| HHS 6 | 2,886 (14.7) | 2,609 (15.0) | 277 (15.6) | 0 (0.0) | 0 (0.0) | 277 (13.1) |
| HHS 7 | 358 (1.8) | 302 (1.7) | 52 (2.9) | 4 (1.4) | 0 (0.0) | 56 (2.6) |
| HHS 8 | 19 (0.1) | 18 (0.1) | 1 (0.1) | 0 (0.0) | 0 (0.0) | 1 (0.0) |
| HHS 9 | 1,561 (8.0) | 1,509 (8.6) | 51 (2.9) | 1 (0.3) | 0 (0.0) | 52 (2.5) |
| HHS 10 | 936 (4.8) | 842 (4.8) | 94 (5.3) | 0 (0.0) | 0 (0.0) | 94 (4.4) |
| Urbanicity^d^, n (column %) |  |  |  |  |  |  |
| City | 7,294 (37.3) | 6,941 (39.8) | 315 (17.8) | 37 (12.8) | 0 (0.0) | 352 (16.6) |
| Suburban | 6,721 (34.3) | 6,137 (35.1) | 506 (28.6) | 78 (26.9) | 5 (8.3) | 589 (27.8) |
| Town | 1,658 (8.5) | 1,305 (7.5) | 274 (15.5) | 63 (21.7) | 12 (20.0) | 349 (16.5) |
| Rural | 3,847 (19.7) | 3,022 (17.3) | 674 (38.1) | 111 (38.3) | 43 (71.7) | 828 (39.1) |
| Unknown | 62 (0.3) | 57 (0.3) | 1 (0.1) | 1 (0.3) | 0 (0.0) | 2 (0.1) |

^a^ PUSC is defined as a school closure lasting ≥5 school days, excluding any scheduled days off.

^b^ Percentages may not add up to 100%, as they are rounded to the nearest tenth of a percent.

^c^ Regions of the United States Department of Health & Human Services (HHS). https://www.hhs.gov/about/agencies/regional-offices/index.html

^d^ Urbanicity was not specified for 5 schools with NCES IDs, and unknown for 57 schools without NCES IDs.

S3 Table. Cause subcategories of prolonged unplanned school closures (PUSCs) by academic year, United States, 2011–2019^a,b^

|  | PUSC by Academic Year | | | | | | | | |
| --- | --- | --- | --- | --- | --- | --- | --- | --- | --- |
|  | Total | 2011-12 | 2012-13 | 2013-14 | 2014-15 | 2015-16 | 2016-17 | 2017-18 | 2018-19 |
| Total, n (row%*) | 22,112 | 770  (3.5) | 4,513 (20.4) | 996 (4.5) | 1,498 (6.8) | 2,495 (11.3) | 1,382 (6.3) | 7,215 (32.6) | 3,243 (14.7) |
| Cause of PUSC^a^, n (column %*) |  | | | | | | | | |
| Weather |  |  |  |  |  |  |  |  |  |
| Ice/snow/cold | 6,823  (87.8) | 623  (100.0) | 186  (42.9) | 854  (100.0) | 1,418  (100.0) | 1,712  (81.4) | 85  (24.2) | 493  (92.8) | 1,452  (99.0) |
| Rain (heavy/severe/tropical  storms) | 947  (12.2) | 0  (0.0) | 248  (57.1) | 0  (0.0) | 0  (0.0) | 380  (18.2) | 266  (75.8) | 38  (7.2) | 15  (1.0) |
| Natural disaster |  |  |  |  |  |  |  |  |  |
| Hurricane | 9,554  (91.0) | 5  (38.5) | 3,407  (99.9) | 0  (0.0) | 0  (0.0) | 267  (85.9) | 844  (86.0) | 3,936  (91.1) | 1,095  (77.2) |
| Wildfire | 621  (5.9) | 0  (0.0) | 3  (0.1) | 0  (0.0) | 1  (8.3) | 35 (11.3) | 10  (1.0) | 382 (8.8) | 190  (13.4) |
| Flood (river/creek) | 111  (1.1) | 0  (0.0) | 0  (0.0) | 6  (19.4) | 0  (0.0) | 8  (2.6) | 77  (7.9) | 1  (0.0) | 19  (1.3) |
| Tornado | 103  (1.0) | 8  (61.5) | 0  (0.0) | 25  (80.7) | 5  (41.7) | 1  (0.3) | 50  (5.1) | 0  (0.0) | 14  (1.0) |
| Earthquake | 101  (1.0) | 0  (0.0) | 0  (0.0) | 0  (0.0) | 0  (0.0) | 0  (0.0) | 0  (0.0) | 1  (0.0) | 100  (7.1) |
| Volcanic eruption | 6  (0.1) | 0  (0.0) | 0  (0.0) | 0  (0.0) | 6  (50.0) | 0  (0.0) | 0  (0.0) | 0  (0.0) | 0  (0.0) |
| Budget/teacher strike |  |  |  |  |  |  |  |  |  |
| Teacher strike | 3,263  (99.9) | 97  (100.0) | 649  (99.4) | 0  (0.0) | 21  (100.0) | 61  (100.0) | 9  (100.0) | 2,239  (100.0) | 183  (100.0) |
| No state funding | 4  (0.1) | 0  (0.0) | 4  (0.6) | 0  (0.0) | 0  (0.0) | 0  (0.0) | 0  (0.0) | 0  (0.0) | 0  (0.0) |
| Environmental problem |  |  |  |  |  |  |  |  |  |
| Asbestos | 3  (1.5) | 0  (0.0) | 0  (0.0) | 0  (0.0) | 3  (100.0) | 0  (0.0) | 0  (0.0) | 0  (0.0) | 0  (0.0) |
| Lead | 3  (1.5) | 0  (0.0) | 0  (0.0) | 0  (0.0) | 0  (0.0) | 3  (21.4) | 0  (0.0) | 0  (0.0) | 0  (0.0) |
| Mold | 74  (36.5) | 4  (100.0) | 6 (60.0) | 1  (1.1) | 0  (0.0) | 11  (78.6) | 3  (100.0) | 1  (3.5) | 48  (100.0) |
| Water contaminant | 91  (44.8) | 0  (0.0) | 0 (0.0) | 91  (98.9) | 0  (0.0) | 0  (0.0) | 0  (0.0) | 0  (0.0) | 0  (0.0) |
| Poor air quality | 32  (15.8) | 0  (0.0) | 4 (40.0) | 0  (0.0) | 0  (0.0) | 0  (0.0) | 0  (0.0) | 28  (96.6) | 0  (0.0) |
| Building/utility problem |  |  |  |  |  |  |  |  |  |
| Building issue^c^ | 52  (46.0) | 5  (15.2) | 3  (50.0) | 1  (11.1) | 11  (64.7) | 8  (66.7) | 6  (50.0) | 8  (80.0) | 10  (71.4) |
| Facilities issue^d^ | 61  (54.0) | 28  (84.9) | 3  (50.0) | 8  (88.9) | 6  (35.3) | 4  (33.3) | 6  (50.0) | 2  (20.0) | 4  (28.6) |
| Illness |  |  |  |  |  |  |  |  |  |
| Influenza/influenza-like  illness | 129  (56.3) | 0  (0.0) | 0  (0.0) | 0  (0.0) | 0  (0.0) | 0  (0.0) | 0  (0.0) | 58  (68.2) | 71  (63.4) |
| Other respiratory illness | 26  (11.4) | 0  (0.0) | 0  (0.0) | 0  (0.0) | 0  (0.0) | 0  (0.0) | 26  (100.0) | 0  (0.0) | 0  (0.0) |
| Gastrointestinal illness | 12  (5.2) | 0  (0.0) | 0  (0.0) | 3  (100.0) | 0  (0.0) | 1  (100.0) | 0  (0.0) | 4  (4.7) | 4  (3.6) |
| Meningitis | 3  (1.3) | 0  (0.0) | 0  (0.0) | 0  (0.0) | 0  (0.0) | 0  (0.0) | 0  (0.0) | 0  (0.0) | 3  (2.7) |
| Unknown illness | 59  (25.8) | 0  (0.0) | 0  (0.0) | 0  (0.0) | 2  (100.0) | 0  (0.0) | 0  (0.0) | 23  (27.1) | 34  (30.4) |
| Violence |  |  |  |  |  |  |  |  |  |
| Actualized violence | 9  (23.7) | 0  (0.0) | 0  (0.0) | 7  (100.0) | 1  (4.0) | 0  (0.0) | 0  (0.0) | 1  (100.0) | 0  (0.0) |
| Violence in the community  – safety precaution | 28  (73.7) | 0  (0.0) | 0  (0.0) | 0  (0.0) | 24  (96.0) | 4  (100.0) | 0  (0.0) | 0  (0.0) | 0  (0.0) |
| Threat | 1  (2.6) | 0  (0.0) | 0  (0.0) | 0  (0.0) | 0  (0.0) | 0  (0.0) | 0  (0.0) | 0  (0.0) | 1  (100.0) |

^a^ PUSC is defined as a school closure lasting ≥5 school days, excluding any scheduled days off.

^b^ Percentages may not add up to 100%, as they are rounded to the nearest tenth of a percent.

^c^ Includes building damage from storm, fire in the building, flood from broken pipe, gas leak, unsafe building structure, vandalism/robbery,

rat and roach infestation, ventilation issue.

^d^ Includes facility issues, no water/unsafe water, no air conditioning, no heat, no electricity.

S4 Table. Causes of prolonged unplanned school closures ^a^ (PUSCs) by academic year and HHS region^b^, United States, 2011–2019^c^

|  | Total^d^  n (column %) | HHS Regions^e^ | | | | | | | | | |
| --- | --- | --- | --- | --- | --- | --- | --- | --- | --- | --- | --- |
|  |  | HHS 1 | HHS 2 | HHS 3 | HHS 4 | HHS 5 | HHS 6 | HHS 7 | HHS 8 | HHS 9 | HHS 10 |
| Total | 22,112 | 1,021 (4.6) | 3,150 (14.3) | 3,109 (14.1) | 6,944 (31.4) | 1,643 (7.4) | 3,143 (14.3) | 418 (1.9) | 20 (0.1) | 1,614 (7.3) | 1,030 (4.7) |
| Cause of PUSC | | | | | | | | | | | |
| Weather | 7,770 | 712 (9.2) | 107 (1.4) | 2,100 (27.0) | 2,028 (26.1) | 916 (11.8) | 982 (12.6) | 326 (4.2) | 0 (0.0) | 10 (0.1) | 589 (7.6) |
| By academic year |  |  |  |  |  |  |  |  |  |  |  |
| 2011-12 | 623 (8.0) | 474 (76.1) | 24 (3.9) | 0 (0.0) | 0 (0.0) | 0 (0.0) | 0 (0.0) | 0 (0.0) | 0 (0.0) | 0 (0.0) | 125 (20.1) |
| 2012-13 | 434 (5.6) | 159 (36.6) | 0 (0.0) | 0 (0.0) | 13 (3.0) | 4 (0.9) | 254 (58.5) | 4 (0.9) | 0 (0.0) | 0 (0.0) | 0 (0.0) |
| 2013-14 | 854 (11.0) | 0 (0.0) | 1 (0.1) | 39 (4.6) | 294 (34.4) | 166 (19.4) | 187 (21.9) | 146 (17.1) | 0 (0.0) | 0 (0.0) | 21 (2.5) |
| 2014-15 | 1,418 (18.3) | 45 (3.2) | 82 (5.8) | 411 (29.0) | 814 (57.4) | 20 (1.4) | 1 (0.1) | 45 (3.2) | 0 (0.0) | 0 (0.0) | 0 (0.0) |
| 2015-16 | 2,092 (26.9) | 0 (0.0) | 0 (0.0) | 1,392 (66.5) | 314 (15.0) | 6 (0.3) | 321 (15.3) | 0 (0.0) | 0 (0.0) | 1 (0.1) | 58 (2.8) |
| 2016-17 | 351 (4.5) | 0 (0.0) | 0 (0.0) | 11 (3.1) | 4 (1.1) | 0 (0.0) | 211 (60.1) | 33 (9.4) | 0 (0.0) | 7 (2.0) | 85 (24.2) |
| 2017-18 | 531 (6.8) | 34 (6.4) | 0 (0.0) | 131 (24.7) | 229 (43.1) | 132 (24.9) | 2 (0.4) | 0 (0.0) | 0 (0.0) | 0 (0.0) | 3 (0.6) |
| 2018-19 | 1,467 (18.9) | 0 (0.0) | 0 (0.0) | 116 (7.9) | 360 (24.5) | 588 (40.1) | 6 (0.4) | 98 (6.7) | 0 (0.0) | 2 (0.1) | 297 (20.3) |
| Natural disaster | 10,496 | 298 (2.8) | 3,018 (28.8) | 115 (1.1) | 4,709 (44.9) | 29 (0.3) | 1,492 (14.2) | 54 (0.5) | 6 (0.1) | 672 (6.4) | 103 (1.0) |
| By academic year |  |  |  |  |  |  |  |  |  |  |  |
| 2011-12 | 13 (0.1) | 1 (7.7) | 3 (23.1) | 1 (7.7) | 6 (46.2) | 2 (15.4) | 0 (0.0) | 0 (0.0) | 0 (0.0) | 0 (0.0) | 0 (0.0) |
| 2012-13 | 3,410 (32.5) | 297 (8.7) | 3,015 (88.4) | 85 (2.5) | 10 (0.3) | 0 (0.0) | 0 (0.0) | 0 (0.0) | 0 (0.0) | 0 (0.0) | 3 (0.1) |
| 2013-14 | 31 (0.3) | 0 (0.0) | 0 (0.0) | 0 (0.0) | 0 (0.0) | 21 (67.7) | 0 (0.0) | 4 (12.9) | 6 (19.4) | 0 (0.0) | 0 (0.0) |
| 2014-15 | 12 (0.1) | 0 (0.0) | 0 (0.0) | 0 (0.0) | 0 (0.0) | 0 (0.0) | 5 (41.7) | 0 (0.0) | 0 (0.0) | 7 (58.3) | 0 (0.0) |
| 2015-16 | 311 (3.0) | 0 (0.0) | 0 (0.0) | 0 (0.0) | 267 (85.9) | 6 (1.9) | 3 (1.0) | 0 (0.0) | 0 (0.0) | 35 (11.3) | 0 (0.0) |
| 2016-17 | 981 (9.4) | 0 (0.0) | 0 (0.0) | 0 (0.0) | 890 (90.7) | 0 (0.0) | 4 (0.4) | 31 (3.2) | 0 (0.0) | 56 (5.7) | 0 (0.0) |
| 2017-18 | 4,320 (41.2) | 0 (0.0) | 0 (0.0) | 0 (0.0) | 2,457 (56.9) | 0 (0.0) | 1,480 (34.3) | 0 (0.0) | 0 (0.0) | 383 (8.9) | 0 (0.0) |
| 2018-19 | 1,418 (13.5) | 0 (0.0) | 0 (0.0) | 29 (2.1) | 1,079 (76.1) | 0 (0.0) | 0 (0.0) | 19 (1.3) | 0 (0.0) | 191 (13.4) | 100 (7.1) |
| Budget/teacher strike | 3,263 | 0 (0.0) | 0 (0.0) | 716 (21.9) | 0 (0.0) | 690 (21.2) | 654 (20.0) | 0 (0.0) | 0 (0.0) | 910 (27.9) | 293 (9.0) |
| By academic year |  |  |  |  |  |  |  |  |  |  |  |
| 2011-12 | 97 (3.0) | 0 (0.0) | 0 (0.0) | 1 (1.0) | 0 (0.0) | 0 (0.0) | 0 (0.0) | 0 (0.0) | 0 (0.0) | 0 (0.0) | 96 (99.0) |
| 2012-13 | 653 (20.0) | 0 (0.0) | 0 (0.0) | 0 (0.0) | 0 (0.0) | 653 (100.0) | 0 (0.0) | 0 (0.0) | 0 (0.0) | 0 (0.0) | 0 (0.0) |
| 2013-14 | 0 (0.0) | 0 (0.0) | 0 (0.0) | 0 (0.0) | 0 (0.0) | 0 (0.0) | 0 (0.0) | 0 (0.0) | 0 (0.0) | 0 (0.0) | 0 (0.0) |
| 2014-15 | 21 (0.6) | 0 (0.0) | 0 (0.0) | 0 (0.0) | 0 (0.0) | 21 (100.0) | 0 (0.0) | 0 (0.0) | 0 (0.0) | 0 (0.0) | 0 (0.0) |
| 2015-16 | 61 (1.9) | 0 (0.0) | 0 (0.0) | 21 (34.4) | 0 (0.0) | 6 (9.8) | 0 (0.0) | 0 (0.0) | 0 (0.0) | 0 (0.0) | 34 (55.7) |
| 2016-17 | 9 (0.3) | 0 (0.0) | 0 (0.0) | 9 (100.0) | 0 (0.0) | 0 (0.0) | 0 (0.0) | 0 (0.0) | 0 (0.0) | 0 (0.0) | 0 (0.0) |
| 2017-18 | 2,239 (68.6) | 0 (0.0) | 0 (0.0) | 675 (30.2) | 0 (0.0) | 0 (0.0) | 654 (29.2) | 0 (0.0) | 0 (0.0) | 910 (40.6) | 0 (0.0) |
| 2018-19 | 183 (5.6) | 0 (0.0) | 0 (0.0) | 10 (5.5) | 0 (0.0) | 10 (5.5) | 0 (0.0) | 0 (0.0) | 0 (0.0) | 0 (0.0) | 163 (89.1) |
| Environmental problem | 203 | 2 (1.0) | 20 (9.9) | 156 (76.9) | 3 (1.5) | 4 (2.0) | 1 (0.5) | 1 (0.5) | 1 (0.5) | 13 (6.4) | 2 (1.0) |
| By academic year |  |  |  |  |  |  |  |  |  |  |  |
| 2011-12 | 4 (2.0) | 0 (0.0) | 3 (75.0) | 0 (0.0) | 0 (0.0) | 0 (0.0) | 0 (0.0) | 1 (25.0) | 0 (0.0) | 0 (0.0) | 0 (0.0) |
| 2012-13 | 10 (4.9) | 0 (0.0) | 7 (70.0) | 0 (0.0) | 1 (10.0) | 0 (0.0) | 1 (10.0) | 0 (0.0) | 0 (0.0) | 0 (0.0) | 1 (10.0) |
| 2013-14 | 92 (45.3) | 0 (0.0) | 0 (0.0) | 91 (98.9) | 0 (0.0) | 1 (1.1) | 0 (0.0) | 0 (0.0) | 0 (0.0) | 0 (0.0) | 0 (0.0) |
| 2014-15 | 3 (1.5) | 0 (0.0) | 0 (0.0) | 0 (0.0) | 0 (0.0) | 0 (0.0) | 0 (0.0) | 0 (0.0) | 0 (0.0) | 3 (100.0) | 0 (0.0) |
| 2015-16 | 14 (6.9) | 0 (0.0) | 0 (0.0) | 1 (7.1) | 1 (7.1) | 3 (21.4) | 0 (0.0) | 0 (0.0) | 0 (0.0) | 9 (64.3) | 0 (0.0) |
| 2016-17 | 3 (1.5) | 0 (0.0) | 0 (0.0) | 0 (0.0) | 0 (0.0) | 0 (0.0) | 0 (0.0) | 0 (0.0) | 1 (33.3) | 1 (33.3) | 1 (33.3) |
| 2017-18 | 29 (14.3) | 0 (0.0) | 0 (0.0) | 28 (96.6) | 1 (3.5) | 0 (0.0) | 0 (0.0) | 0 (0.0) | 0 (0.0) | 0 (0.0) | 0 (0.0) |
| 2018-19 | 48 (23.7) | 2 (4.2) | 10 (20.8) | 36 (75.0) | 0 (0.0) | 0 (0.0) | 0 (0.0) | 0 (0.0) | 0 (0.0) | 0 (0.0) | 0 (0.0) |
| Building/utility problem | 113 | 9 (8.0) | 5 (4.4) | 16 (14.2) | 3 (2.7) | 2 (1.8) | 22 (19.5) | 3 (2.7) | 12 (10.6) | 4 (3.5) | 37 (32.7) |
| By academic year |  |  |  |  |  |  |  |  |  |  |  |
| 2011-12 | 33 (29.2) | 0 (0.0) | 0 (0.0) | 0 (0.0) | 0 (0.0) | 0 (0.0) | 0 (0.0) | 0 (0.0) | 0 (0.0) | 1 (3.0) | 32 (97.0) |
| 2012-13 | 6 (5.3) | 3 (50.0) | 0 (0.0) | 0 (0.0) | 0 (0.0) | 0 (0.0) | 0 (0.0) | 0 (0.0) | 0 (0.0) | 0 (0.0) | 3 (50.0) |
| 2013-14 | 9 (8.0) | 1 (11.1) | 0 (0.0) | 3 (33.3) | 0 (0.0) | 0 (0.0) | 0 (0.0) | 0 (0.0) | 5 (55.6) | 0 (0.0) | 0 (0.0) |
| 2014-15 | 17 (15.0) | 1 (5.9) | 1 (5.9) | 4 (23.5) | 0 (0.0) | 1 (5.9) | 4 (23.5) | 3 (17.7) | 3 (17.7) | 0 (0.0) | 0 (0.0) |
| 2015-16 | 12 (10.6) | 0 (0.0) | 1 (8.3) | 4 (33.3) | 0 (0.0) | 1 (8.3) | 5 (41.7) | 0 (0.0) | 0 (0.0) | 0 (0.0) | 1 (8.3) |
| 2016-17 | 12 (10.6) | 2 (16.7) | 1 (8.3) | 0 (0.0) | 0 (0.0) | 0 (0.0) | 5 (41.7) | 0 (0.0) | 2 (16.7) | 2 (16.7) | 0 (0.0) |
| 2017-18 | 10 (8.9) | 0 (0.0) | 2 (20.0) | 1 (10.0) | 0 (0.0) | 0 (0.0) | 5 (50.0) | 0 (0.0) | 0 (0.0) | 1 (10.0) | 1 (10.0) |
| 2018-19 | 14 (12.4) | 2 (14.3) | 0 (0.0) | 4 (28.6) | 3 (21.4) | 0 (0.0) | 3 (21.4) | 0 (0.0) | 2 (14.3) | 0 (0.0) | 0 (0.0) |
| Illness | 229 | 0 (0.0) | 0 (0.0) | 0 (0.0) | 200 (87.3) | 2 (0.9) | 11 (4.8) | 11 (4.8) | 0 (0.0) | 4 (1.8) | 1 (0.4) |
| By academic year |  |  |  |  |  |  |  |  |  |  |  |
| 2011-12 | 0 (0.0) | 0 (0.0) | 0 (0.0) | 0 (0.0) | 0 (0.0) | 0 (0.0) | 0 (0.0) | 0 (0.0) | 0 (0.0) | 0 (0.0) | 0 (0.0) |
| 2012-13 | 0 (0.0) | 0 (0.0) | 0 (0.0) | 0 (0.0) | 0 (0.0) | 0 (0.0) | 0 (0.0) | 0 (0.0) | 0 (0.0) | 0 (0.0) | 0 (0.0) |
| 2013-14 | 3 (1.3) | 0 (0.0) | 0 (0.0) | 0 (0.0) | 0 (0.0) | 0 (0.0) | 3 (100.0) | 0 (0.0) | 0 (0.0) | 0 (0.0) | 0 (0.0) |
| 2014-15 | 2 (0.9) | 0 (0.0) | 0 (0.0) | 0 (0.0) | 2 (100.0) | 0 (0.0) | 0 (0.0) | 0 (0.0) | 0 (0.0) | 0 (0.0) | 0 (0.0) |
| 2015-16 | 1 (0.4) | 0 (0.0) | 0 (0.0) | 0 (0.0) | 1 (100.0) | 0 (0.0) | 0 (0.0) | 0 (0.0) | 0 (0.0) | 0 (0.0) | 0 (0.0) |
| 2016-17 | 26 (11.4) | 0 (0.0) | 0 (0.0) | 0 (0.0) | 26 (100.0) | 0 (0.0) | 0 (0.0) | 0 (0.0) | 0 (0.0) | 0 (0.0) | 0 (0.0) |
| 2017-18 | 85 (37.1) | 0 (0.0) | 0 (0.0) | 0 (0.0) | 77 (90.6) | 0 (0.0) | 4 (4.7) | 0 (0.0) | 0 (0.0) | 4 (4.7) | 0 (0.0) |
| 2018-19 | 112 (48.9) | 0 (0.0) | 0 (0.0) | 0 (0.0) | 94 (83.9) | 2 (1.8) | 4 (3.6) | 11 (9.8) | 0 (0.0) | 0 (0.0) | 1 (0.9) |
| Violence | 38 | 0 (0.0) | 0 (0.0) | 6 (15.8) | 1 (2.6) | 0 (0.0) | 1 (2.6) | 23 (60.5) | 1 (2.6) | 1 (2.6) | 5 (13.2) |
| By academic year |  |  |  |  |  |  |  |  |  |  |  |
| 2011-12 | 0 (0.0) | 0 (0.0) | 0 (0.0) | 0 (0.0) | 0 (0.0) | 0 (0.0) | 0 (0.0) | 0 (0.0) | 0 (0.0) | 0 (0.0) | 0 (0.0) |
| 2012-13 | 0 (0.0) | 0 (0.0) | 0 (0.0) | 0 (0.0) | 0 (0.0) | 0 (0.0) | 0 (0.0) | 0 (0.0) | 0 (0.0) | 0 (0.0) | 0 (0.0) |
| 2013-14 | 7 (18.4) | 0 (0.0) | 0 (0.0) | 5 (71.4) | 0 (0.0) | 0 (0.0) | 0 (0.0) | 0 (0.0) | 1 (14.3) | 1 (14.3) | 0 (0.0) |
| 2014-15 | 25 (65.8) | 0 (0.0) | 0 (0.0) | 1 (4.0) | 0 (0.0) | 0 (0.0) | 0 (0.0) | 23 (92.0) | 0 (0.0) | 0 (0.0) | 1 (4.0) |
| 2015-16 | 4 (10.5) | 0 (0.0) | 0 (0.0) | 0 (0.0) | 0 (0.0) | 0 (0.0) | 0 (0.0) | 0 (0.0) | 0 (0.0) | 0 (0.0) | 4 (100.0) |
| 2016-17 | 0 (0.0) | 0 (0.0) | 0 (0.0) | 0 (0.0) | 0 (0.0) | 0 (0.0) | 0 (0.0) | 0 (0.0) | 0 (0.0) | 0 (0.0) | 0 (0.0) |
| 2017-18 | 1 (2.6) | 0 (0.0) | 0 (0.0) | 0 (0.0) | 1 (100.0) | 0 (0.0) | 0 (0.0) | 0 (0.0) | 0 (0.0) | 0 (0.0) | 0 (0.0) |
| 2018-19 | 1 (2.6) | 0 (0.0) | 0 (0.0) | 0 (0.0) | 0 (0.0) | 0 (0.0) | 1 (100.0) | 0 (0.0) | 0 (0.0) | 0 (0.0) | 0 (0.0) |

^a^ PUSC is defined as a school closure lasting ≥5 school days, excluding any scheduled days off.

^b^ Regions of the United States Department of Health & Human Services (HHS). https://www.hhs.gov/about/agencies/regional-offices/index.html

^c^ Percentages may not add up to 100%, as they are rounded to the nearest tenth of a percent.

^d^ n (column percentages).

^e^ n (row percentages).

S5 Table. Top ten states with natural disaster-related prolonged unplanned school closures (PUSCs ) by type of natural disaster, United States, 2011–2019^a,b^.

|  | Total | Cause of PUSC (column %) | | | | | |
| --- | --- | --- | --- | --- | --- | --- | --- |
|  |  | Hurricane | Wildfire | Flood | Tornado | Earthquake | Volcanic eruption |
| Total, n (row %) | 10,496 | 9,554 (91.0) | 621 (5.9) | 111 (1.1) | 103 (1.0) | 101 (1.0) | 6 (0.1) |
| Top ten states,  n (column %) |  |  |  |  |  |  |  |
| Florida | 2,409 (23.0) | 2,409 (25.2) | 0 (0.0) | 0 (0.0) | 0 (0.0) | 0 (0.0) | 0 (0.0) |
| New York | 1.997 (19.0) | 1,997 (21.0) | 0 (0.0) | 0 (0.0) | 0 (0.0) | 0 (0.0) | 0 (0.0) |
| Texas | 1,491 (14.2) | 1,480 (15.5) | 0 (0.0) | 2 (1.8) | 9 (8.7) | 0 (0.0) | 0 (0.0) |
| North Carolina | 1,106 (10.5) | 1,106 (11.6) | 0 (0.0) | 0 (0.0) | 0 (0.0) | 0 (0.0) | 0 (0.0) |
| New Jersey | 1,021 (9.7) | 1,021 (10.7) | 0 (0.0) | 0 (0.0) | 0 (0.0) | 0 (0.0) | 0 (0.0) |
| South Carolina | 776 (7.4) | 776 (8.1) | 0 (0.0) | 0 (0.0) | 0 (0.0) | 0 (0.0) | 0 (0.0) |
| California | 663 (6.3) | 0 (0.0) | 618 (99.5) | 45 (40.5) | 0 (0.0) | 0 (0.0) | 0 (0.0) |
| Georgia | 380 (3.6) | 334 (3.5) | 0 (0.0) | 0 (0.0) | 46 (44.7) | 0 (0.0) | 0 (0.0) |
| Connecticut | 297 (2.8) | 297 (3.1) | 0 (0.0) | 0 (0.0) | 0 (0.0) | 0 (0.0) | 0 (0.0) |
| Alaska | 100 (1.0) | 0 (0.0) | 0 (0.0) | 0 (0.0) | 0 (0.0) | 100 (99.0) | 0 (0.0) |

^a^ PUSC is defined as a school closure lasting ≥5 school days, excluding any scheduled days off.

^b^ An additional 17 states experienced a total of 256 (2.4%) natural disaster-related PUSCs and percentages are rounded to the nearest tenth of a percent, therefore percentages may not add up to 100%.

S6 Table. Seasonality of weather-related, prolonged unplanned school closures ^a^ (PUSCs) by HHS Region^b^, United States, 2011–2019^c^

|  | Total | HHS Regions | | | | | | | | | |
| --- | --- | --- | --- | --- | --- | --- | --- | --- | --- | --- | --- |
|  |  | HHS 1 | HHS 2 | HHS 3 | HHS 4 | HHS 5 | HHS 6 | HHS 7 | HHS 8 | HHS 9 | HHS 10 |
| Total weather-related PUSCs, n (row %) | 7,770 | 712 (9.2) | 107 (1.4) | 2,100 (27.0) | 2,028 (26.1) | 916 (11.8) | 982 (12.6) | 326 (4.2) | 0  (0.0) | 10 (0.1) | 589 (7.6) |
| Season of weather-related PUSCs, n (column %) |  |  |  |  |  |  |  |  |  |  |  |
| Fall | 711 (9.2) | 496 (69.7) | 106 (99.1) | 1  (0.1) | 0  (0.0) | 4  (0.4) | 8  (0.8) | 31  (9.5) | 0  (0.0) | 7  (70.0) | 58  (9.9) |
| Winter | 6,060  (78.0) | 204  (28.7) | 1  (0.9) | 2,088  (99.4) | 1,967  (97.0) | 908  (99.1) | 199  (20.3) | 284  (87.1) | 0  (0.0) | 2  (20.0) | 407  (69.1) |
| Spring | 523  (6.7) | 12  (1.7) | 0  (0.0) | 0  (0.0) | 60  (3.0) | 4  (0.4) | 321  (32.7) | 9  (2.8) | 0  (0.0) | 1  (10.0) | 116  (19.7) |
| Summer | 476  (6.1) | 0  (0.0) | 0  (0.0) | 11  (0.5) | 1  (0.1) | 0  (0.0) | 454  (46.2) | 2  (0.6) | 0  (0.0) | 0  (0.0) | 8  (1.4) |

^a^ PUSC is defined as a school closure lasting ≥5 school days, excluding any scheduled days off.

^b^ Regions of the United States Department of Health & Human Services (HHS). https://www.hhs.gov/about/agencies/regional-offices/index.html

^C^ Percentages may not add up to 100%, as they are rounded to the nearest tenth of a percent.

S7 Table. States with illness-related prolonged unplanned school closures (PUSCs ) by illness category, United States, 2011–2019^a,b^.

|  | Total | Cause of PUSC | | | |
| --- | --- | --- | --- | --- | --- |
|  |  | Influenza/influenza- like illness | Gastrointestinal illness | Meningitis | Unknown illness^c^ |
| Total, n (row %) | 229 | 155 (67.7) | 12 (5.2) | 3 (1.3) | 59 (25.8) |
| State, n (column %) |  |  |  |  |  |
| Kentucky | 120 (52.4) | 95 (61.3) | 0 (0.0) | 0 (0.0) | 25 (42.4) |
| Tennessee | 78 (34.1) | 44 (28.4) | 0 (0.0) | 0 (0.0) | 34 (57.6) |
| Texas | 8 (3.5) | 5 (3.2) | 0 (0.0) | 3 (100.0) | 0 (0.0) |
| Missouri | 7 (3.1) | 7 (4.5) | 0 (0.0) | 0 (0.0) | 0 (0.0) |
| California | 4 (1.8) | 0 (0.0) | 4 (33.3) | 0 (0.0) | 0 (0.0) |
| Iowa | 4 (1.8) | 0 (0.0) | 4 (33.3) | 0 (0.0) | 0 (0.0) |
| Oklahoma | 3 (1.3) | 0 (0.0) | 3 (25.0) | 0 (0.0) | 0 (0.0) |
| Alabama | 2 (0.9) | 1 (0.7) | 1 (8.3) | 0 (0.0) | 0 (0.0) |
| Minnesota | 2 (0.9) | 2 (1.3) | 0 (0.0) | 0 (0.0) | 0 (0.0) |
| Idaho | 1 (0.4) | 1 (0.7) | 0 (0.0) | 0 (0.0) | 0 (0.0) |

^a^ PUSC is defined as a school closure lasting ≥5 school days, excluding any scheduled days off.

^b^ Percentages may not add up to 100%, as they are rounded to the nearest tenth of a percent.

^c^ The closure announcement did not specify the type(s) of illness.
